# BOADICEA model: updates to the *BRCA2* breast cancer risks for ages 60 years and older

**DOI:** 10.1101/2024.04.18.24305970

**Authors:** Lorenzo Ficorella, Xin Yang, Douglas F Easton, Antonis C Antoniou

## Abstract

Breast cancer risks in older *BRCA2* pathogenic variant carriers are understudied. Recent studies show a marked decline in the relative risk at older ages. We used data from two large studies to update the breast cancer risks in the BOADICEA model for *BRCA2* carriers 60 years and older.

## Main Text

The Breast and Ovarian Analysis of Disease Incidence and Carrier Estimation Algorithm (BOADICEA), implemented in the CanRisk tool (www.canrisk.org) [1], can be used to predict the risk of developing breast cancer (BC) and the likelihood of carrying pathogenic variants in breast cancer susceptibility genes [2, 3]. The model combines the effects of rare genetic variants, polygenic risk scores, cancer family history, mammographic density, and several other lifestyle/hormonal risk factors.

The risks associated with *BRCA1* and *BRCA2* pathogenic variants (PVs) used in BOADICEA were originally estimated using complex segregation analysis in a large series of families carrying PVs. The model estimated relative risks (RRs) of breast and ovarian cancer up to age 70 years, as there were few data at older ages [4]. The model assumed breast cancer log-relative risks that are piecewise linear functions of age: in particular, the RRs were estimated to increase from 8.99 at age 59 to 13.1 at age 69 years. To implement the model for older individuals, the RR was assumed to remain constant at 13.1 for ages 70 years and older [2, 4]. However, more recent data [5, 6, 7] suggest that the BC RRs decrease with age, and that the RR for ages 70 and over is much lower; therefore, the original RR estimate of 13.1 is most likely an overestimate for older *BRCA2* carriers.

We therefore updated the RRs in BOADICEA by re-deriving the piecewise log-RR linear function for ages 60 and over. We first estimated a revised RR for ages 70-79 using results from two recent large studies [5, 6]. The first, BRIDGES, is a case-control analysis based on panel testing in ∼50,000 breast cancer cases and 50,000 controls from population-based studies; this study showed a clear decline in the odds ratio (OR) by age and reported an OR of 3.05 [95% CI: 2.14-4.35] at ages 60-80. Another large case-control study did not report age-specific ORs, but also found a decline with age; inspection of the results presented graphically show OR at ages 70-80 of ∼3 [7]. The second study is a prospective cohort study that included 1,610 *BRCA2* PV carriers unaffected at baseline; this study reported a RR for ages 71-80 of 6. 6 [95% CI: 3.0-14.7]. We derived a meta-analysis RR estimate for ages 70 and over (3.38) as the inverse-variance weighted average of these two estimates (using the reported sample sizes and 95% confidence intervals to compute the variances). To ensure that relative risks decrease smoothly with age, a piece-wise log-linear function with age was then fitted such that the BC RR decreases from 9.05 at age 60 (as in the previous model) to 3.38 at ages 70, and constant thereafter. Note that the RRs here correspond to the average RR for *BRCA2* carriers relative to the population incidences, averaged over all polygenic effects, which are the inputs to the model [2]. The residual polygenic component in the model was adjusted to account for the change in the RRs and to ensure that the model remains internally consistent for predicting the breast cancer familial relative risks [3].

Figure 1 shows the revised age-dependent BC RRs for *BRCA2* PV carriers (panel A), and the predicted cumulative risk of developing BC for *BRCA2* PV carriers (panels B, C). Based on the revised RRs, the predicted cumulative risk of developing BC for *BRCA2* PV carriers with unknown family history (panel B) remains identical to the previous BOADICEA model until around age 61, and increases more slowly thereafter, reaching 58% by age 80. This compares to the 77% BC risk by age 80 obtained in the previous version. For a *BRCA2* PV carrier with an affected mother at age 40 (panel C), the breast cancer risk by age 80 is predicted to be 72.5%, compared to the 87% BC risk obtained in the previous version. These revised predictions are more consistent with both the population-based penetrance estimates (e.g. ∼43% in the BRIDGES study [5], ∼50% reported by Hu *et al*. [7]) and estimates from carriers with cancer family history (72% [95% CI: 65%-79%] reported by Kuchenbaecker *et al*. [6]).

**Figure 1.**
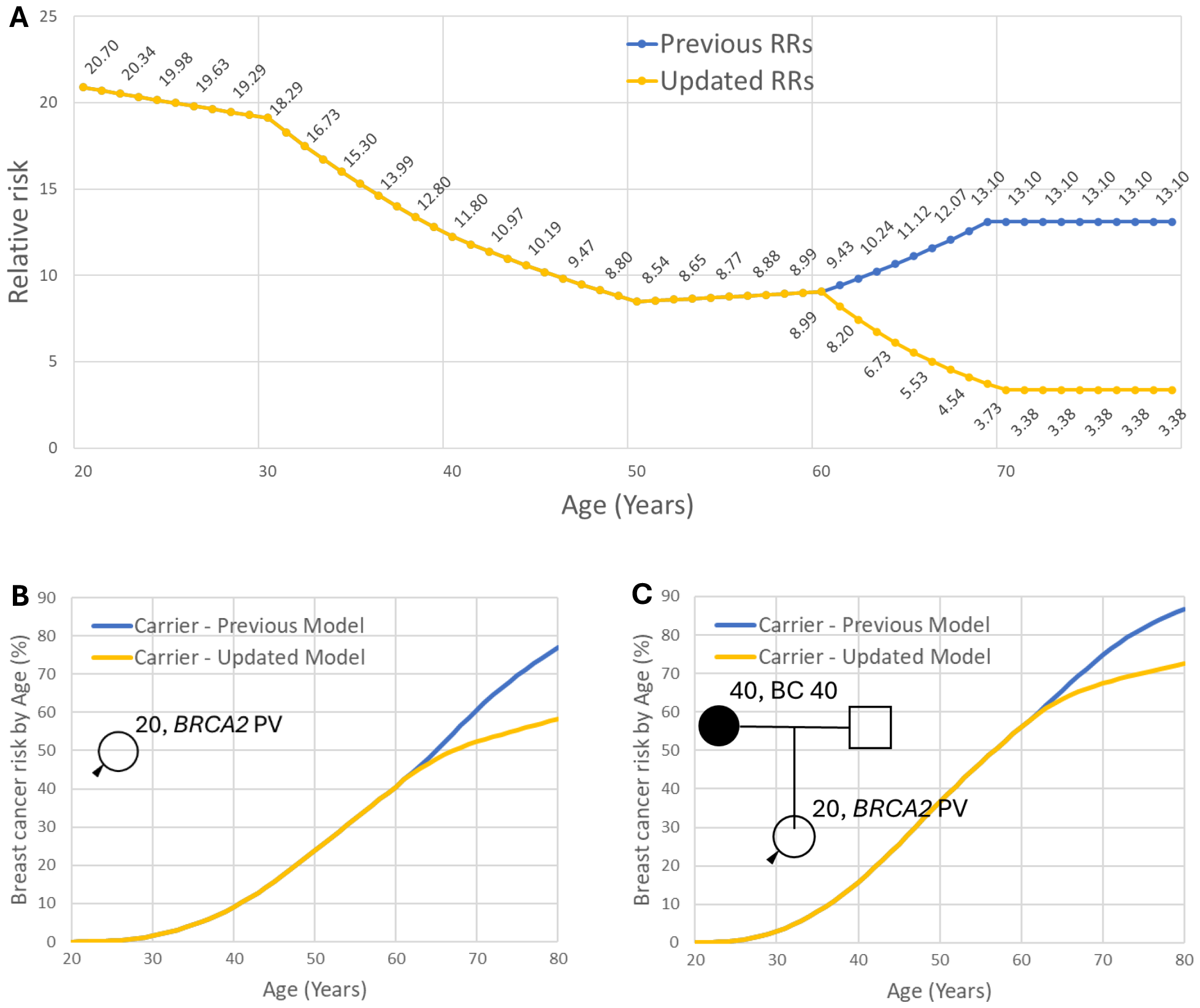
Panel A: previous and updated *BRCA2* relative risks in BOADICEA/CanRisk v2.4. Panel B: revised cumulative breast cancer risk for the average *BRCA2* pathogenic variant (PV) carrier (20y old, born in 2000) using the updated *BRCA2* RRs. Panel C: Corresponding cumulative risks for *BRCA2* PV carrier (20y old, born in 2000) with an affected mother (40y old, born in 1980) diagnosed at age 40.

We note that the data are still limited in this age-group and that there is still uncertainty in the risk estimates, and that better estimates may be become available with further data. The estimates from the cohort studies are likely to be upwardly biased (as estimates of the RRs relative to the general population) as the cohort is oversampled for carriers with a family history. However, the BRIDGES study (though large) has relatively few control carriers, and we did not want to rely solely on the results from one study. All studies, however, show a consistent decline in the RR with age (a feature which would be appear also more plausible mechanistically that the previous increase) and the new estimates are therefore likely to be more realistic. A recent validation study in *BRCA1* and *BRCA2* PV carriers suggests that this model provides calibrated risks in both *BRCA2* PV carriers under and over age 50 years old [8]. The updated RRs are also included in the corresponding ovarian cancer risk model [9]. The updated breast and ovarian cancer models are available through the CanRisk v2.4 tool.

## Data Availability

All data produced in the present work are contained in the manuscript (or referenced literature)

## Author declarations

### Contributions

Conceptualisation: LF, XY, DFE, ACA; Analysis & Model development: LF; Writing in initial draft: LF, DFE, ACA. All authors reviewed the manuscript, provided feedback and approved the final manuscript text.

## Acknowledgements

This work was supported by Cancer Research UK grant: PPRPGM-Nov20\100002

## References

1. Carver T, Hartley S, Lee A, Cunningham AP, Archer S, Babb de Villiers C, Roberts J, Ruston R, Walter FM, Tischkowitz M, Easton DF, Antoniou AC. CanRisk Tool-A Web Interface for the Prediction of Breast and Ovarian Cancer Risk and the Likelihood of Carrying Genetic Pathogenic Variants. Cancer Epidemiol Biomarkers Prev. 2021 Mar;30(3):469–473. doi: 10.1158/1055-9965.EPI-20-1319.

2. Lee A, Mavaddat N, Cunningham A, Carver T, Ficorella L, Archer S, Walter FM, Tischkowitz M, Roberts J, Usher-Smith J, Simard J, Schmidt MK, Devilee P, Zadnik V, Jurgens H, Mouret-Fourme E, De Pauw A, Rookus M, Mooij TM, Pharoah PP, Easton DF, Antoniou AC. Enhancing the BOADICEA cancer risk prediction model to incorporate new data on RAD51C, RAD51D, BARD1 updates to tumour pathology and cancer incidence. J Med Genet 2022;59(12):1206–18 doi: 10.1136/jmedgenet-2022-108471

3. Lee A, Mavaddat N, Wilcox AN, Cunningham AP, Carver T, Hartley S, Babb de Villiers C, Izquierdo A, Simard J, Schmidt MK, Walter FM, Chatterjee N, Garcia-Closas M, Tischkowitz M, Pharoah P, Easton DF, Antoniou AC. BOADICEA: a comprehensive breast cancer risk prediction model incorporating genetic and nongenetic risk factors. Genet Med. 2019 Aug;21(8):1708–1718. doi: 10.1038/s41436-018-0406-9

4. Antoniou AC, Cunningham AP, Peto J, Evans DG, Lalloo F, Narod SA, Risch HA, Eyfjord JE, Hopper JL, Southey MC, Olsson H, Johannsson O, Borg A, Pasini B, Radice P, Manoukian S, Eccles DM, Tang N, Olah E, Anton-Culver H, Warner E, Lubinski J, Gronwald J, Gorski B, Tryggvadottir L, Syrjakoski K, Kallioniemi OP, Eerola H, Nevanlinna H, Pharoah PD, Easton DF. The BOADICEA model of genetic susceptibility to breast and ovarian cancers: updates and extensions. Br J Cancer 2008;98(8):1457–66 doi: 10.1038/sj.bjc.6604305

5. Breast Cancer Association C, Dorling L, Carvalho S, Allen J, Gonzalez-Neira A, Luccarini C, Wahlstrom C, Pooley KA, Parsons MT, Fortuno C, Wang Q, Bolla MK, Dennis J, Keeman R, Alonso MR, Alvarez N, Herraez B, Fernandez V, Nunez-Torres R, Osorio A, Valcich J, Li M, Torngren T, Harrington PA, Baynes C, Conroy DM, Decker B, Fachal L, Mavaddat N, Ahearn T, Aittomaki K, Antonenkova NN, Arnold N, Arveux P, Ausems M, Auvinen P, Becher H, Beckmann MW, Behrens S, Bermisheva M, Bialkowska K, Blomqvist C, Bogdanova NV, Bogdanova-Markov N, Bojesen SE, Bonanni B, Borresen-Dale AL, Brauch H, Bremer M, Briceno I, Bruning T, Burwinkel B, Cameron DA, Camp NJ, Campbell A, Carracedo A, Castelao JE, Cessna MH, Chanock SJ, Christiansen H, Collee JM, Cordina-Duverger E, Cornelissen S, Czene K, Dork T, Ekici AB, Engel C, Eriksson M, Fasching PA, Figueroa J, Flyger H, Forsti A, Gabrielson M, Gago-Dominguez M, Georgoulias V, Gil F, Giles GG, Glendon G, Garcia EBG, Alnaes GIG, Guenel P, Hadjisavvas A, Haeberle L, Hahnen E, Hall P, Hamann U, Harkness EF, Hartikainen JM, Hartman M, He W, Heemskerk-Gerritsen BAM, Hillemanns P, Hogervorst FBL, Hollestelle A, Ho WK, Hooning MJ, Howell A, Humphreys K, Idris F, Jakubowska A, Jung A, Kapoor PM, Kerin MJ, Khusnutdinova E, Kim SW, Ko YD, Kosma VM, Kristensen VN, Kyriacou K, Lakeman IMM, Lee JW, Lee MH, Li J, Lindblom A, Lo WY, Loizidou MA, Lophatananon A, Lubinski J, MacInnis RJ, Madsen MJ, Mannermaa A, Manoochehri M, Manoukian S, Margolin S, Martinez ME, Maurer T, Mavroudis D, McLean C, Meindl A, Mensenkamp AR, Michailidou K, Miller N, Mohd Taib NA, Muir K, Mulligan AM, Nevanlinna H, Newman WG, Nordestgaard BG, Ng PS, Oosterwijk JC, Park SK, Park-Simon TW, Perez JIA, Peterlongo P, Porteous DJ, Prajzendanc K, Prokofyeva D, Radice P, Rashid MU, Rhenius V, Rookus MA, Rudiger T, Saloustros E, Sawyer EJ, Schmutzler RK, Schneeweiss A, Schurmann P, Shah M, Sohn C, Southey MC, Surowy H, Suvanto M, Thanasitthichai S, Tomlinson I, Torres D, Truong T, Tzardi M, Valova Y, van Asperen CJ, Van Dam RM, van den Ouweland AMW, van der Kolk LE, van Veen EM, Wendt C, Williams JA, Yang XR, Yoon SY, Zamora MP, Evans DG, de la Hoya M, Simard J, Antoniou AC, Borg A, Andrulis IL, Chang-Claude J, Garcia-Closas M, Chenevix-Trench G, Milne RL, Pharoah PDP, Schmidt MK, Spurdle AB, Vreeswijk MPG, Benitez J, Dunning AM, Kvist A, Teo SH, Devilee P, Easton DF. Breast Cancer Risk Genes - Association Analysis in More than 113,000 Women. N Engl J Med 2021;384(5):428–39 doi: 10.1056/NEJMoa1913948

6. Kuchenbaecker KB, Hopper JL, Barnes DR, Phillips KA, Mooij TM, Roos-Blom MJ, Jervis S, van Leeuwen FE, Milne RL, Andrieu N, Goldgar DE, Terry MB, Rookus MA, Easton DF, Antoniou AC, Brca Consortium BC, McGuffog L, Evans DG, Barrowdale D, Frost D, Adlard J, Ong KR, Izatt L, Tischkowitz M, Eeles R, Davidson R, Hodgson S, Ellis S, Nogues C, Lasset C, Stoppa-Lyonnet D, Fricker JP, Faivre L, Berthet P, Hooning MJ, van der Kolk LE, Kets CM, Adank MA, John EM, Chung WK, Andrulis IL, Southey M, Daly MB, Buys SS, Osorio A, Engel C, Kast K, Schmutzler RK, Caldes T, Jakubowska A, Simard J, Friedlander ML, McLachlan SA, Machackova E, Foretova L, Tan YY, Singer CF, Olah E, Gerdes AM, Arver B, Olsson H. Risks of Breast, Ovarian, and Contralateral Breast Cancer for BRCA1 and BRCA2 Mutation Carriers. JAMA 2017;317(23):2402–16 doi: 10.1001/jama.2017.7112

7. Hu C, Hart SN, Gnanaolivu R, Huang H, Lee KY, Na J, Gao C, Lilyquist J, Yadav S, Boddicker NJ, Samara R, Klebba J, Ambrosone CB, Anton-Culver H, Auer P, Bandera EV, Bernstein L, Bertrand KA, Burnside ES, Carter BD, Eliassen H, Gapstur SM, Gaudet M, Haiman C, Hodge JM, Hunter DJ, Jacobs EJ, John EM, Kooperberg C, Kurian AW, Le Marchand L, Lindstroem S, Lindstrom T, Ma H, Neuhausen S, Newcomb PA, O’Brien KM, Olson JE, Ong IM, Pal T, Palmer JR, Patel AV, Reid S, Rosenberg L, Sandler DP, Scott C, Tamimi R, Taylor JA, Trentham-Dietz A, Vachon CM, Weinberg C, Yao S, Ziogas A, Weitzel JN, Goldgar DE, Domchek SM, Nathanson KL, Kraft P, Polley EC, Couch FJ. A Population-Based Study of Genes Previously Implicated in Breast Cancer. N Engl J Med. 2021 Feb 4;384(5):440–451. doi: 10.1056/NEJMoa2005936

8. Yang X, Mooij TM, Leslie G, Ficorella L, Andrieu N, Kast K, Singer CF, Jakubowska A, van Gils CH, Tan Y, Engel C, Adank MA, van Asperen CJ, Ausems Mgem, Berthet P, Embrace Collée JM, Cook J, Eason J, van Spaendonck-Zwarts KY, Evans DG, Gomez Garcia EB, Hanson H, Izatt L, Kemp Z, Lalloo F, Lasset C, Lesueur F, Musgrave H, Nambot S, Nogues C, Oosterwijk JC, Stoppa-Lyonnet D, Tischkowitz M, Tripathi V, Wevers MR, Zhao E, van Leeuwen F, Schmidt MK, Easton DF, Rookus MA, Antoniou AC. Validation of the BOADICEA Model in a Prospective Cohort of BRCA1/2 Pathogenic Carriers. medRxiv [Preprint] 2024. doi: 10.1101/2024.04.21.24306136 [Accessed 22rd April 2024]

9. Lee A, Yang X, Tyrer J, Gentry-Maharaj A, Ryan A, Mavaddat N, Cunningham AP, Carver T, Archer S, Leslie G, Kalsi J, Gaba F, Manchanda R, Gayther S, Ramus SJ, Walter FM, Tischkowitz M, Jacobs I, Menon U, Easton DF, Pharoah P, Antoniou AC. Comprehensive epithelial tubo-ovarian cancer risk prediction model incorporating genetic and epidemiological risk factors. J Med Genet. 2022 Jul;59(7):632–643. doi: 10.1136/jmedgenet-2021-107904

